# A Python Toolkit for Simulated Fall Risk Assessment Using Synthetic Wearable Sensor Data

**DOI:** 10.1101/2025.08.22.25334265

**Authors:** Daniel Opoku-Gyamfi

## Abstract

**Background:** Falls are a leading cause of injury and reduced mobility, particularly among prosthetic users, older adults, and individuals with neuromuscular impairments. Accurate assessment of fall risk is essential for timely interventions, personalized rehabilitation, and overall safety. However, real-world data collection is often constrained by ethical, logistical, and safety considerations, limiting the availability of sufficiently large and diverse datasets.

**Aim:** To develop a synthetic dataset representing low, medium, and high fall risk scenarios and evaluate the performance of machine learning models trained on this data to provide a reproducible framework for fall risk prediction while mitigating challenges associated with real-world data.

**Methods:** A Python-based toolkit was used to generate synthetic accelerometer (accel_x, accel_y, accel_z) and gyroscope (gyro_x, gyro_y, gyro_z) signals for 1,000 samples across three fall risk categories. Key features, including mean, standard deviation, and variability measures, were extracted from these signals. A Random Forest classifier with 100 decision trees was trained on 80% of the dataset and tested on the remaining 20%. Performance was assessed using accuracy, precision, recall, F1-score, and confusion matrices.

**Results:** The classifier achieved an overall accuracy of 80%, with high precision and recall for low-risk (precision 0.90, recall 0.88) and high-risk (precision 0.75, recall 0.88) categories. Medium-risk predictions were less accurate (precision 0.59, recall 0.54). Feature distributions across fall risk levels demonstrated meaningful separation, supporting the utility of synthetic signals in model training.

**Conclusion:** Synthetic accelerometer and gyroscope data can effectively support fall risk classification, offering a reproducible and ethical alternative for algorithm development in the absence of large-scale real-world datasets.

**Clinical Relevance:** This approach facilitates the rapid development and testing of predictive models for fall prevention, enabling safer, data-driven strategies for vulnerable populations such as older adults and prosthetic users.

## Introduction

Falls are a leading cause of injury and reduced mobility, particularly among prosthetic users, older adults, and individuals with neuromuscular impairments (1). Accurate assessment of fall risk is critical for implementing timely interventions, designing personalized rehabilitation programs, and improving overall safety and quality of life (2). Traditional approaches to fall risk assessment rely on wearable sensors, clinical tests, or observational methods; however, collecting real-world data can be challenging (3). Ethical considerations, patient safety, limited access to specialized populations, and logistical constraints often restrict the availability of sufficiently large and diverse datasets for research and algorithm development (4).

Synthetic data provides a promising solution to these challenges by enabling the generation of reproducible, controlled datasets that mimic real-world sensor measurements (5,6). Researchers can develop and validate predictive models without exposing human participants to potential harm by simulating accelerometer and gyroscope signals representative of various fall risk scenarios (7,8). Such synthetic datasets are particularly valuable for methodological development, proof-of-concept studies, and educational purposes, allowing for reproducible and transparent evaluation of machine learning algorithms in fall risk prediction (9,10).

Moreover, synthetic data can be used to address common limitations in real-world datasets, such as class imbalance and underrepresentation of high-risk populations, which often bias model performance and reduce generalizability (11,12). Researchers can create balanced datasets that capture the full spectrum of fall risk, improving the robustness and reliability of predictive models by simulating diverse movement patterns and risk scenarios (13,14). Advances in machine learning and artificial intelligence further enable the integration of synthetic data with real-world measurements, allowing hybrid datasets that leverage the advantages of both approaches to optimize model training and validation (15,16). Additionally, the use of synthetic data facilitates rapid prototyping and testing of novel algorithms, accelerates methodological innovation, and supports reproducible research practices that are critical for advancing fall risk assessment technologies (17–19).

Finally, the application of synthetic datasets extends beyond methodological development. They offer opportunities to evaluate new sensor configurations, optimize feature extraction methods, and explore the impact of individualized rehabilitation strategies in silico before implementation in clinical or home-based environments (20–22). Synthetic datasets help overcome ethical and logistical barriers while still providing realistic and actionable insights for fall prevention, ultimately contributing to safer and more effective interventions for vulnerable populations by reducing reliance on large-scale human data collection (23,24).

In spite of these advances and the potential benefits of synthetic data, there remains a need to systematically evaluate its utility for fall risk prediction using wearable sensor signals, particularly accelerometer and gyroscope data. Accordingly, this study aims to develop a synthetic dataset representing low, medium, and high fall risk scenarios and to assess the performance of machine learning models trained on this data, providing a reproducible framework for advancing fall risk assessment methodologies while mitigating the ethical, logistical, and safety challenges associated with real-world data collection.

## Methods

This study presents a Python-based toolkit for simulating wearable sensor data and predicting fall risk categories. All analyses were implemented in Python (v3.10) using numpy, pandas, scikit-learn, matplotlib, and seaborn libraries.

### Synthetic Data Generation

Synthetic accelerometer (accel_x, accel_y, accel_z) and gyroscope (gyro_x, gyro_y, gyro_z) signals were generated to simulate daily movements representative of low, medium, and high fall risk scenarios, following approaches described in previous studies (25,26). Accelerometer data incorporated variations in movement amplitude and directional changes, while gyroscope data captured angular velocities around each axis. Random Gaussian noise was added to mimic realistic sensor variability. The dataset consisted of 1,000 samples, with class labels assigned based on predefined thresholds for fall risk, resulting in a moderate class imbalance.

### Feature Extraction

Key features were computed from the synthetic signals, including mean, standard deviation, and variability measures for each acceleration and angular velocity component. These features are commonly used to assess postural stability and movement irregularities relevant to fall risk.

### Machine Learning Model

A Random Forest classifier was employed to predict fall risk categories (Low, Medium, High) based on the extracted features. The model used 100 decision trees with a maximum depth of 5, chosen to balance model performance and computational efficiency. A fixed random seed ensured reproducibility, and the dataset was randomly split into 80% training and 20% testing sets, stratified by class to preserve class distributions.

### Evaluation Metrics

Model performance was assessed using multiple metrics: accuracy (overall correct predictions), precision (proportion of correct positive predictions per class), recall (proportion of actual positives correctly identified), and F1-score (harmonic mean of precision and recall). A confusion matrix was also generated to visualize predicted versus actual fall risk distributions. To evaluate model stability, experiments were repeated across multiple random splits.

### Code Availability

The code used to generate the synthetic dataset, train the Random Forest classifier, and reproduce all analyses is publicly available at https://doi.org/10.5281/zenodo.16928374.

All analyses were conducted using Python (v3.10) on Google Colab, which provides a standardized and reproducible computational environment. The use of a fixed random seed, along with versioned libraries, ensures that the synthetic data generation, feature extraction, and model training can be consistently reproduced across different systems.

## Results

The synthetic dataset consisted of accelerometer (accel_x, accel_y, accel_z) and gyroscope (gyro_x, gyro_y, gyro_z) signals with corresponding fall risk labels (Low, Medium, High). A preview of the generated data is shown in Table 1, illustrating the variability across sensor axes and the distribution of fall risk labels in the first few samples.

**Table 1.** Example of synthetic sensor data and fall risk labels.

| accel_x | accel_y | accel_z | gyro_x | gyro_y | gyro_z | fall_risk |
| --- | --- | --- | --- | --- | --- | --- |
| 0.938 | -0.278 | 8.721 | 0.033 | -0.129 | -0.107 | Low |
| 0.518 | 0.736 | 9.259 | 0.135 | -0.029 | 0.204 | Medium |
| 0.109 | -0.713 | 10.821 | -0.001 | -0.074 | 0.016 | Low |
| 2.507 | 2.027 | 9.530 | 0.006 | 0.076 | 0.011 | Medium |
| 1.642 | 0.072 | 9.992 | 0.055 | -0.069 | 0.072 | Low |

Feature distributions across fall risk categories were visualized to explore patterns in accelerometer and gyroscope signals (Figure 1). These plots demonstrate observable differences in signal magnitudes and variability among Low, Medium, and High fall risk classes, suggesting that the extracted features provide meaningful information for classification.

**Figure 1.**
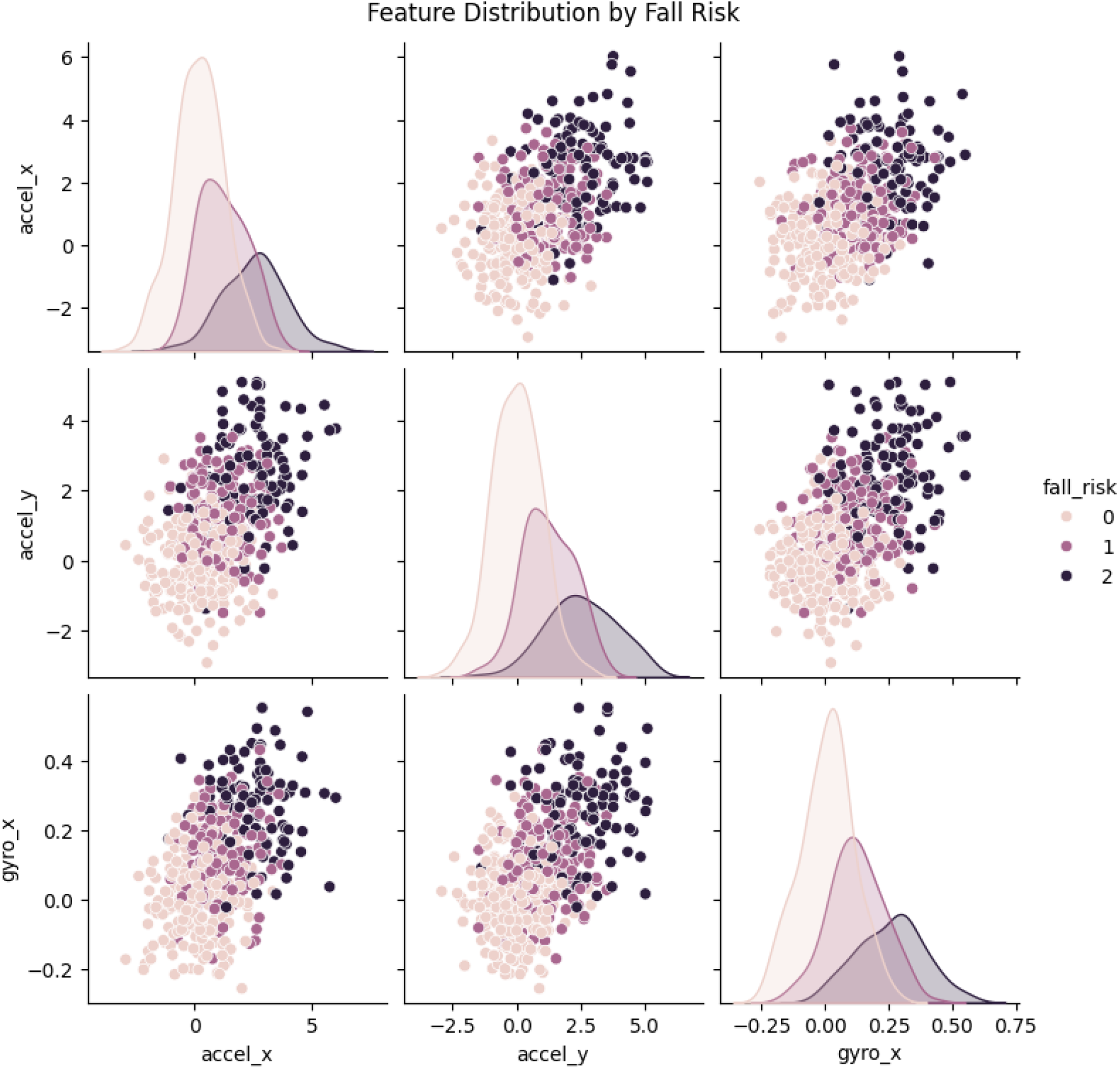
Feature distribution by fall risk

The Random Forest classifier achieved an **overall accuracy of 80%** on the test set. Performance metrics for each fall risk category are presented in the classification report (Table 2). The model performed best in predicting Low-risk cases, with a precision of 0.90, recall of 0.88, and F1-score of 0.89. High-risk predictions also showed strong performance (precision 0.75, recall 0.88, F1-score 0.81). Medium-risk cases were predicted less accurately, with a precision of 0.59, recall of 0.54, and F1-score of 0.57. Macro-averaged metrics indicate balanced overall performance (precision 0.75, recall 0.77, F1-score 0.75), while weighted averages reflect the class distribution in the test set (Table 2).

**Table 2.** Classification performance metrics by fall risk category.

| Class | Precision | Recall | F1-score | Support |
| --- | --- | --- | --- | --- |
| Low | 0.90 | 0.88 | 0.89 | 59 |
| Medium | 0.59 | 0.54 | 0.57 | 24 |
| High | 0.75 | 0.88 | 0.81 | 17 |
| Accuracy |  |  | 0.80 | 100 |
| Macro avg | 0.75 | 0.77 | 0.75 | 100 |
| Weighted avg | 0.80 | 0.80 | 0.80 | 100 |

The confusion matrix further illustrates the classifier’s performance (Figure 2). Most misclassifications occurred between Low and Medium risk categories, consistent with the lower precision and recall observed for the Medium class.

**Figure 2.**
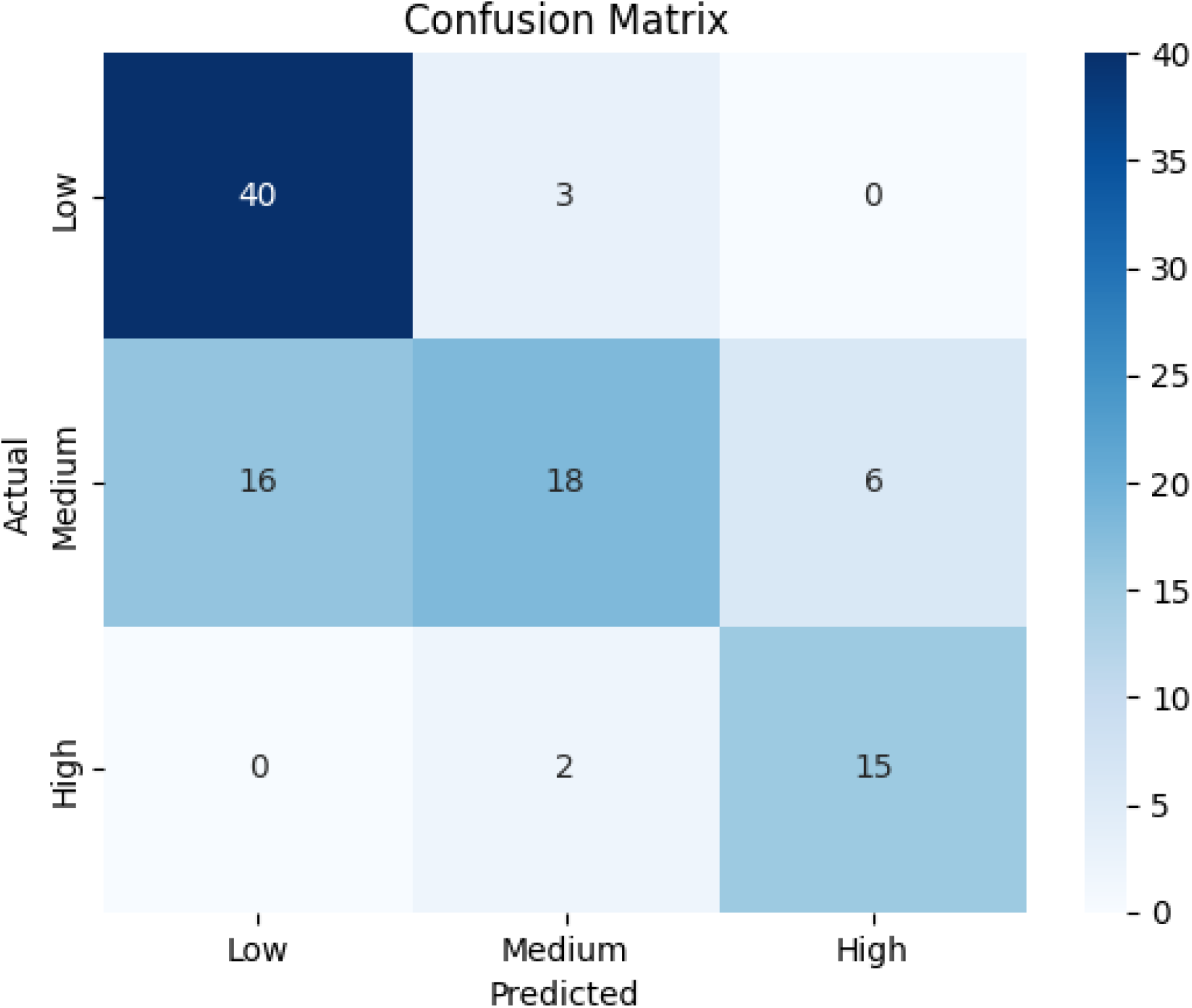
Confusion matrix showing predicted vs. actual fall risk.

These results indicate that the Random Forest model effectively distinguishes between fall risk levels based on the synthetic sensor features, particularly for Low and High risk categories.

## Discussion

This study developed a synthetic dataset of accelerometer and gyroscope signals to simulate low, medium, and high fall risk scenarios, achieving an overall classification accuracy of 80% using a Random Forest classifier. The model demonstrated high precision and recall for low- and high-risk categories, with precision values of 0.90 and 0.75, respectively, highlighting the potential of synthetic data in training predictive models for fall risk assessment.

Recent research has highlighted the efficacy of synthetic data in enhancing machine learning models for fall detection. For instance, Debnath et al. (2025)(27) demonstrated that incorporating diffusion-based synthetic data improved the offline F1-score by 7–10% and boosted real-time fall detection performance by 24%, confirming its value in improving model robustness in real-world settings. Similarly, Lim et al. (2024)(28) explored the use of AI for fall risk prediction, leveraging gait analysis via computer vision and machine learning, emphasizing the importance of diverse and high-quality data sources. Our results also align with studies exploring synthetic fall data generated through Large Language Models and diffusion-based methods, which showed that the quality and characteristics of synthetic datasets critically influence model performance, particularly when real-world fall recordings are scarce (29). Furthermore, work on wrist-based fall detection demonstrated that conditional diffusion-based synthetic data effectively mitigates class imbalance and data scarcity, enhancing model performance even with limited real-world recordings (30). Supporting these findings, clinical studies have shown that synthetic data, including diffusion-based augmentation, can substantially improve fall detection models, echoing reported F1-score gains of 7–10% offline and 24% in real-time (31).

Our findings are also consistent with research using single gait cycle analysis and gradient boosting decision trees, demonstrating that even limited or short-duration sensor data, such as our synthetic accelerometer and gyroscope signals, can effectively classify fall risk, offering accessible, low-burden monitoring solutions (32). In a broader health data science context, synthetic datasets can improve data availability, strengthen model robustness, and address scarcity challenges, paralleling their applications in health economics and outcomes research (33). Additionally, studies employing LLM-generated and diffusion-based synthetic fall data have highlighted that dataset characteristics and sampling frequency critically affect model performance, particularly when real-world fall recordings are limited (29). Finally, research integrating accelerometric and non-accelerometric data supports the notion that combining multiple data sources improves fall risk prediction accuracy, consistent with the superior performance of Bayesian ridge regression in older adult cohorts (34).

Wearable sensor systems for fall risk assessment, particularly inertial measurement units (IMUs), have been extensively reviewed and shown to provide meaningful gait information essential for predicting falls (35). Our synthetic dataset mirrors these real-world sensor outputs, offering a reproducible and ethical resource for developing and testing predictive models. Mohan et al. (2025)(36) developed a cooperative AI-based meta-model combining Fuzzy Logic and Deep Belief Networks to predict fall risk, achieving 90% overall accuracy, 100% specificity, and 85.71% sensitivity by leveraging vital signs and daily activity patterns for personalized prediction. In comparison, our study used a Random Forest classifier on fully synthetic sensor data, achieving 80% accuracy. While medium-risk predictions were comparatively lower in our model, both approaches underscore the utility of machine learning in fall risk assessment, with Mohan et al. focusing on real-world physiological and activity data, and our study demonstrating the value of synthetic data for reproducible model development.

Several limitations should be noted. The synthetic dataset, while useful for initial model development, may not capture the full complexity and variability of real-world scenarios. As Mohan et al. (2025)(36) observed, synthetic data can impair model generalizability and increase overfitting risks, particularly when datasets are small or non-homogeneous. Moreover, the class imbalance in our dataset, with more low-risk samples, may have influenced performance metrics. Future studies should integrate real-world datasets to validate and refine models developed using synthetic data.

This study demonstrates the feasibility of using synthetic accelerometer and gyroscope data for fall risk classification. Our findings provide a foundation for future work aimed at integrating real-world data to enhance model accuracy, generalizability, and clinical applicability across diverse populations.

## Conclusion

This study demonstrates that synthetic accelerometer and gyroscope data can be effectively used to classify fall risk into low, medium, and high categories, achieving meaningful predictive performance with a Random Forest classifier. The results underscore the potential of synthetic datasets to support the development and testing of machine learning models in scenarios where real-world data are scarce or difficult to collect, offering a reproducible and ethical alternative for methodological research. While limitations related to dataset complexity, class imbalance, and generalizability remain, the study establishes a foundation for integrating synthetic and real-world data to enhance model accuracy and applicability. Future research should focus on validating these models with diverse populations and real-world recordings to further improve fall risk prediction and enable practical deployment in clinical and home-based monitoring systems.

## Data Availability

All data produced are available online at https://doi.org/10.5281/zenodo.16928374

## Declarations

### Author Contribution

Daniel Opoku-Gyamfi conceived the study, developed the methodology and Python code, generated the synthetic dataset, performed data analysis, interpreted the results, and wrote the manuscript.

### Ethics Approval and Consent to Participate

Not applicable, as this study exclusively used synthetic data and did not involve human participants.

### Consent for Publication

Not applicable.

### Data and Code Availability

The synthetic dataset and all Python code used for data generation, feature extraction, model training, and evaluation are publicly available at https://doi.org/10.5281/zenodo.16928374

## References

1. Hunter SW, Batchelor F, Hill KD, Hill AM, Mackintosh S, Payne M. Risk Factors for Falls in People With a Lower Limb Amputation: A Systematic Review. PM&R [Internet]. 2017;9(2):170-180.e1. Available from: https://www.sciencedirect.com/science/article/pii/S1934148216308140

2. Miura T, Kanoya Y. Fall Risk Assessment and Prevention Strategies in Nursing Homes: A Narrative Review. Healthc (Basel, Switzerland). 2025 Feb;13(4).

3. Ferreira RN, Ribeiro NF, Santos CP. Fall Risk Assessment Using Wearable Sensors: A Narrative Review. Vol. 22, Sensors. 2022.

4. McCradden MD, Joshi S, Mazwi M, Anderson JA. Ethical limitations of algorithmic fairness solutions in health care machine learning. Lancet Digit Heal [Internet]. 2020 May 1;2(5):e221–3. Available from: 10.1016/S2589-7500(20)30065-0

5. Marwala T. Synthetic Versus Authentic Data BT - The Balancing Problem in the Governance of Artificial Intelligence. In: Marwala T, editor. Singapore: Springer Nature Singapore; 2024. p. 105–20. Available from: 10.1007/978-981-97-9251-1_7

6. Liu F, Panagiotakos D. Real-world data: a brief review of the methods, applications, challenges and opportunities. BMC Med Res Methodol [Internet]. 2022;22(1):287. Available from: 10.1186/s12874-022-01768-6

7. Howcroft J, Kofman J, Lemaire ED. Prospective Fall-Risk Prediction Models for Older Adults Based on Wearable Sensors. IEEE Trans Neural Syst Rehabil Eng. 2017;25(10):1812–20.

8. Subramaniam S, Faisal AI, Deen MJ. Wearable sensor systems for fall risk assessment: A review. Front Digit Heal. 2022;4:921506.

9. Luna-Romero SF, Abreu de Souza M, Serpa Andrade L. Artificial Vision Systems for Mobility Impairment Detection: Integrating Synthetic Data, Ethical Considerations, and Real-World Applications. Vol. 13, Technologies. 2025.

10. Moore J, Stuart S, McMeekin P, Walker R, Nouredanesh M, Tung J, et al. Toward enhanced free-living fall risk assessment: Data mining and deep learning for environment and terrain classification. Intell Med [Internet]. 2023;8:100103. Available from: https://www.sciencedirect.com/science/article/pii/S2666521223000170

11. Uddin MZ. Synthetic Data BT - Trustworthy Multimodal Intelligent Systems for Independent Living. In: Uddin MZ, editor. Cham: Springer Nature Switzerland; 2025. p. 51–109. Available from: 10.1007/978-3-031-97359-8_3

12. Whitney CD, Norman J. Real risks of fake data: Synthetic data, diversity-washing and consent circumvention. In: Proceedings of the 2024 ACM Conference on Fairness, Accountability, and Transparency. 2024. p. 1733–44.

13. Rajagopalan R, Litvan I, Jung TP. Fall Prediction and Prevention Systems: Recent Trends, Challenges, and Future Research Directions. Vol. 17, Sensors. 2017.

14. Bargiotas I, Wang D, Mantilla J, Quijoux F, Moreau A, Vidal C, et al. Preventing falls: the use of machine learning for the prediction of future falls in individuals without history of fall. J Neurol. 2022 Jul 11;270.

15. Alkhalifah T, Wang H, Ovcharenko O. MLReal: Bridging the gap between training on synthetic data and real data applications in machine learning. Artif Intell Geosci [Internet]. 2022;3:101–14. Available from: https://www.sciencedirect.com/science/article/pii/S2666544122000260

16. Kumar A. The Role of Synthetic Data in Advancing AI Models: Opportunities, Challenges, and Ethical Considerations. J Artif Intell Gen Sci ISSN3006-4023 [Internet]. 2024 Aug 7;5(1 SE-Articles):443–59. Available from: https://newjaigs.org/index.php/JAIGS/article/view/256

17. González-Castro A, Leirós-Rodríguez R, Prada-García C, Benítez-Andrades JA. The Applications of Artificial Intelligence for Assessing Fall Risk: Systematic Review. J Med Internet Res [Internet]. 2024;26:e54934. Available from: https://www.jmir.org/2024/1/e54934

18. Stier SP, Kreisbeck C, Ihssen H, Popp MA, Hauch J, Malek K, et al. Materials acceleration platforms (MAPs): accelerating materials research and development to meet urgent societal challenges. Adv Mater. 2024;36(45):2407791.

19. Bouras A. The emerging applications of synthetic data in neurosurgery research and practice: a systematic review. medRxiv. 2024;2003–24.

20. Zaher M, Ghoneim A, Abdelhamid L, Atia A. Artificial intelligence techniques in enhancing home-based rehabilitation: A survey. FCI-H Informatics Bull. 2024;6(2):16–30.

21. Vourganas I, Stankovic V, Stankovic L. Individualised Responsible Artificial Intelligence for Home-Based Rehabilitation. Vol. 21, Sensors. 2021.

22. Boltaboyeva A, Baigarayeva Z, Imanbek B, Ozhikenov K, Getahun AJ, Aidarova T, et al. A Review of Innovative Medical Rehabilitation Systems with Scalable AI-Assisted Platforms for Sensor-Based Recovery Monitoring. Vol. 15, Applied Sciences. 2025.

23. Shanley D, Hogenboom J, Lysen F, Wee L, Lobo Gomes A, Dekker A, et al. Getting real about synthetic data ethics: Are AI ethics principles a good starting point for synthetic data ethics? EMBO Rep. 2024 May;25(5):2152–5.

24. Hao S, Han W, Jiang T, Li Y, Wu H, Zhong C, et al. Synthetic data in AI: Challenges, applications, and ethical implications. arXiv Prepr arXiv240101629. 2024;

25. Lee H. Fall Detection Method based on Deep Learning using Accelerometer and Gyroscope Data [Internet]. 서 울 대 학 교 대 학 원; 2022. Available from: https://s-space.snu.ac.kr/handle/10371/181345

26. Raizman O. Daily Life Activity Recognition with a Head Mounted IMU on Older Adults. 2023; Available from: https://repository.tudelft.nl/file/File_1abf5889-f38f-49cb-b265-89f5ed2573ae?preview=1

27. Debnath M, Alamgeer S, Kabir MS, Ngu AH. Enhancing Wearable Fall Detection System via Synthetic Data. Sensors (Basel). 2025 Jul;25(15).

28. Lim ZK, Connie T, Goh MKO, Saedon N ‘Izzati B. Fall risk prediction using temporal gait features and machine learning approaches. Front Artif Intell. 2024;7:1425713.

29. Alamgeer S, Souissi Y, Ngu A. AI-Generated Fall Data: Assessing LLMs and Diffusion Model for Wearable Fall Detection. Vol. 25, Sensors. 2025.

30. Tu YC, Lin CY, Liu CP, Chan CT. Performance Analysis of Data Augmentation Approaches for Improving Wrist-Based Fall Detection System. Vol. 25, Sensors. 2025.

31. Debnath M, Kabir MS, Ni J, Ngu AHH. The Impact of Synthetic Data on Fall Detection Application BT - Artificial Intelligence in Medicine. In: Finkelstein J, Moskovitch R, Parimbelli E, editors. Cham: Springer Nature Switzerland; 2024. p. 204–9.

32. Nishiyama D, Arita S, Fukui D, Yamanaka M, Yamada H. Accurate fall risk classification in elderly using one gait cycle data and machine learning. Clin Biomech [Internet]. 2024;115:106262. Available from: https://www.sciencedirect.com/science/article/pii/S0268003324000949

33. Lai TC, Ngorsuraches S. Roles of AI-Based Synthetic Data in Health Economics and Outcomes Research. Value Heal [Internet]. 2025; Available from: https://www.sciencedirect.com/science/article/pii/S1098301525023125

34. González-Castro Ana, Benítez-Andrades José Alberto González-González Rubén, Prada-García Camino, Leirós-Rodríguez Raquel. Predicting fall risk in older adults: A machine learning comparison of accelerometric and non-accelerometric factors. Digit Heal [Internet]. 2025 Mar 28;11:20552076251331750. Available from: 10.1177/20552076251331752

35. Capodici A, Fanconi C, Curtin C, Shapiro A, Noci F, Giannoni A, et al. A scoping review of machine learning models to predict risk of falls in elders, without using sensor data. Diagnostic Progn Res. 2025 May;9(1):11.

36. Mohan D, Chong PH, Gutierrez J. A Novel Cooperative AI-Based Fall Risk Prediction Model for Older Adults. Vol. 25, Sensors. 2025.

